# Evaluating the Efficacy of a Hybrid Physical Therapy Technology: A Pilot Study on Wearable Sleeves within a Veteran Patient Population

**DOI:** 10.1101/2025.01.21.25320682

**Authors:** Jeremy Truntzer, Shannon Gorman

## Abstract

**Introduction:** Traditional musculoskeletal rehabilitation presents a range of challenges that can hinder patient recovery. Cost, access and travel place a significant burden on many health care systems. We completed a pilot study using a wearable biosleeve to determine if such technology might help address many of the current challenges.

**Methods:** Patients were enrolled following knee arthroscopy to receive hybrid physical therapy utilizing a wearable sleeve that tracks joint angles and biometrics in conjunction with a web-based platform that allows provider visibility to patient completion data for assigned protocols.

**Results:** Ten patients, who completed a three-month rehabilitation program, experienced successful outcomes with no adverse events. No difference in adverse outcomes or failure to achieve acceptable motion was found in patients that completed hybrid therapy (in-person and digital) or digital-based therapy alone.

**Conclusion:** While larger studies are required, this study serves as an important launching point for the potential benefits of integrating wearable technology in musculoskeletal rehabilitation.

## Introduction

Traditional physical rehabilitation for musculoskeletal care presents a range of challenges that can impact patient outcomes. Such traditional means often requires in-person interaction, either at the home or the outpatient setting, subjecting the process to delays in initiation, high costs, and patient resources in the form of transportation and inconvenience. Further, the accuracy and resolution of tools are inadequate, limiting objective measurement capability.

**Access to therapy** remains perhaps the most critical issue. Availability of physical therapy services can vary significantly, particularly in rural or underserved communities where physical therapists are scarce.^1–3^ This inconsistency in access can lead to disparities in recovery outcomes, as patients may not receive the necessary rehabilitation due to travel or service availability constraints.^4^ Moreover, delay in care is a known contributor to adverse outcomes and preventable factor leading to secondary procedures. Waiting 28 days to see a PT can be detrimental to the short- and long-term outcomes of MSK injuries.^5,6^ Access to board-certified specialist physical therapists is even more limited. When telehealth options are offered to patients, there is variability in what “telehealth” means; for example, over a third of Medicare beneficiaries receiving remote care were only offered a phone visit, even though 65% of those patients were video capable.^7^

The **financial burden** associated with multiple physical therapy sessions can be considerable. Research indicates that the average cost of physical therapy can vary widely, with some sessions ranging from $50 to $350, depending on the facility and geographic location. This variability often restricts access for many patients, particularly those in lower socio-economic brackets.^8,9^ Studies report therapy-associated costs exceeding $2500 following common procedures, with variation depending on insurance provider.^10^ The high costs call into question the adequacy of coverage for therapy despite clear importance.^11^

Additionally, the **travel burden** to therapy appointments can negatively impact patient compliance and engagement. Patients living far from rehabilitation centers often find adhering to a prescribed therapy regimen challenging due to the associated time and costs of travel. Studies have shown that such logistical barriers can result in decreased therapeutic adherence and overall treatment efficacy.^10^ Multiple studies have identified that telehealth visits have a lower no-show rate than in-person visits, suggesting that increasing the availability of telehealth therapy may improve patient adherence to therapy.^12,13^

In response to the numerous challenges in the current physical therapy landscape, providers and patients are turning to emerging technologies and pathways. Wearable devices present an innovative solution to address these challenges by offering customizable rehabilitation programs, remote engagement and objective data feedback to both patient and provider. However, even with newer technology, there remains gaps in measurement capability and patient engagement. This pilot program aimed to assess the initial efficacy and safety of a wearable technology in the post-operative setting that provides cost effective and accurate measurement of motion features such as joint angles and biosignals. The integrated solution was tested within a select Veteran patient population.

## Methods

Veteran patients (n=10) who underwent knee arthroscopy with a single fellowship-trained surgeon were enrolled in this pilot study. This study was approved by the Institutional Review Board and all patients signed a consent prior to enrollment. The participants were enrolled in a post-operative therapy program and equipped with the OpenMotion AI (Saratoga, CA) BioSleeve™ - a flexible garment with a mesh of embedded sensors (**FIGURE 1**). Patients were instructed to wear the sleeve during prescribed rehabilitation exercises only. Initiation of home therapy with the sleeve was started within the first week of therapy. Phased protocols were customized by providers and assigned to patients based on the operative procedure within the Open Motion AI Kaleidoscope™ web-application (**FIGURE 2**). Sleeves tracked motion, biomechanical features such as joint angles, and biosignals such as vital signs – data which are then transmitted to the web-based platform. Patients were monitored via standard clinic visits or telehealth video appointments with the surgeon. Analytics and visualizations were provided to both patients and providers accessible via the application portal. Patients completed at home therapy using the wearable sleeve often along with routine outpatient therapy per prior arrangement. Patient satisfaction and adverse events were recorded.

**FIGURE 1:**
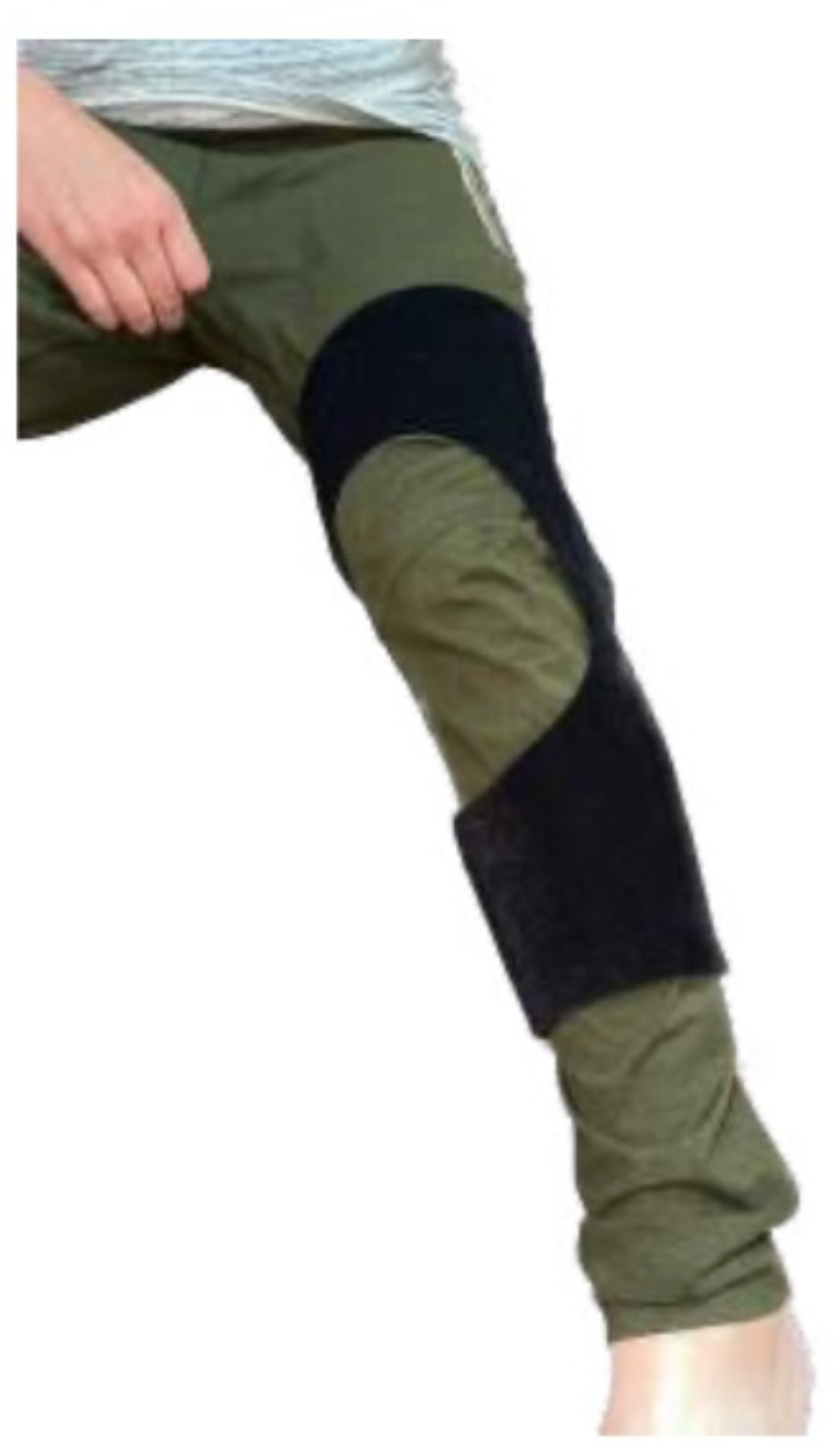
Lower Extremity Open Motion AI Biosleeve.

**FIGURE 2:**
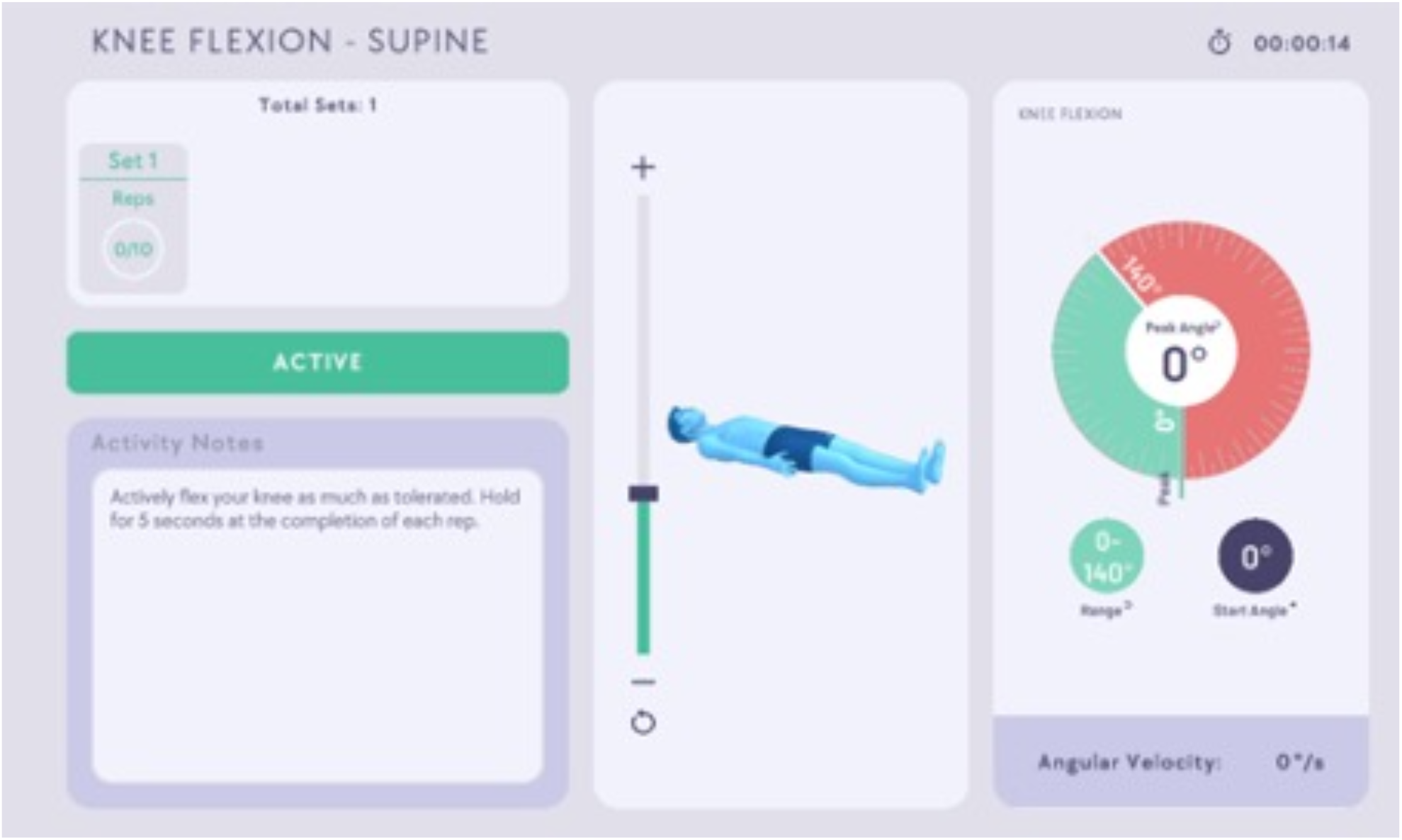
Kaleidoscope web-application interface that pairs with the Open Motion AI Biosleeve.

## Results

A total of ten patients were enrolled in the study and successfully completed a 3-month course. All 10 patients were Veterans, male, aged 36.3 (range 24-64 yrs) (**TABLE 1**). Laterality included six right knees and four left knees. Seven of the patients received outpatient in-person therapy compared with three that engaged in remote digital therapy only using the wearable sleeve. Six of the patients traveled over 90 miles for surgical care, thus completed surgical follow-up care via telehealth video visits.

**TABLE 1:** Patient Characteristics.

| Age | Gender | Laterality | Indication | Procedure | Hybrid or Telehealth* |
| --- | --- | --- | --- | --- | --- |
| 45-49 | Male | Right | Flexion contracture | Arthroscopic lysis of adhesions | Hybrid |
| 40-44 | Male | Right | Meniscus tear | Meniscal debridement | Telehealth |
| 30-34 | Male | Left | Meniscus tear | Meniscal debridement | Telehealth |
| 60-64 | Male | Left | Meniscus tear | Meniscal debridement | Hybrid |
| 25-29 | Male | Right | Meniscus tear | Meniscal debridement | Telehealth |
| 20-24 | Male | Left | Meniscus tear | Meniscal debridement | Telehealth |
| 25-29 | Male | Left | Meniscus tear | Meniscal debridement | Telehealth |
| 35-39 | Male | Right | Cartilage lesion | Chondroplasty | Hybrid |
| 35-39 | Male | Right | Meniscus tear | Meniscal debridement | Telehealth |
| 30-34 | Male | Right | Meniscus tear | Meniscus repair | Hybrid |

\*Patient surgical follow-up was completed via telehealth (digital) or combination of telehealth and in-person evaluation

At the completion of the study all patients obtained full range of motion as determined by exceeding or meeting pre-operative levels. No skin or adverse events were reported related to use of the wearable sleeve. Four patients reported challenges with onboarding due to charging issues of the sleeve that were resolved by the company support team. In all cases, care was not delayed more than 3 days. All patients noted high satisfaction with the technology and would consider using the wearable sleeve in the future.

## Discussion

This pilot study indicates that the utilization of a hybrid therapy model including a wearable sleeve in combination with a digital rehabilitation program may positively impact post- operative recovery from musculoskeletal injury. The absence of adverse events highlights the safety of this technology, while the achievement of full range of motion in all participants suggests its potential efficacy in improving rehabilitation outcomes. Additionally, the model is promising for addressing several of the challenges in the broader context of current physical therapy practices.

Consideration of digital physical therapy has gained traction recently given the multiple limitations of conventional therapy. The Covid pandemic is also responsible for accelerating adoption. Studies have noted high retention rates and patient satisfaction with digital therapy.^14,15^ Digital therapy has proven a viable model for shoulder ailments.^16–18^ Similarly, effective results are reported for telehealth models following total hip and knee arthroplasty.^19–21^

Within the Veteran population specifically, where access barriers to care are well-known, telehealth models have shown great promise for expanding timely musculoskeletal care.^22^ Hatch et al found that success in telehealth utilization increased with rural location, living far away from a VA facility, and high co-payments within a Veteran population.^23^ A study assessing functional mobility in wheeled mobility devices found larger and clinically significant changes in transfer scores with telehealth compared to in-person care.^24^

Although not explored directly in this pilot program, multiple studies have demonstrated the cost-value of digital and telehealth platforms for rehabilitation.^25^ Telehealth-based physical therapy following hip arthroscopy led to significant savings compared to in-person models.^26^ Significant cost savings were also reported for the treatment of chronic knee pain.^27’^

## Conclusion

Overall, digital rehabilitation has proven a viable, effective, and resource saving model across various musculoskeletal domains. Our pilot program provides further evidence for the potential to improve physical therapy via shared objective data, easy engagement, analytics, and telehealth capabilities. Larger studies are needed across various health care models to further evaluate and confirm cost savings, safety, outcomes and generalizability.

## Data Availability

All data produced in the present study are available upon reasonable request to the authors

## Notes

### Competing Interest Statement

The authors have declared no competing interest.

### Funding Statement

This study did not receive any funding

### Author Declarations

IRB of Veterans Affairs, Palo Alto and Stanford University gave ethical approval for this work

